# Evaluation of sampling frequency and normalization of SARS-CoV-2 wastewater concentrations for capturing COVID-19 burdens in the community

**DOI:** 10.1101/2021.02.17.21251867

**Authors:** Shuchen Feng, Adélaïde Roguet, Jill S. McClary-Gutierrez, Ryan J. Newton, Nathan Kloczko, Jonathan G. Meiman, Sandra L. McLellan

## Abstract

Wastewater surveillance for SARS-CoV-2 provides an approach for assessing the infection burden across a city. For these data to be useful for public health, measurement variability and the relationship to case data need to be established. We measured SARS-CoV-2 RNA concentrations in the influent of twelve wastewater treatment plants from August 2020 to January 2021. Replicate samples demonstrated that N1 gene target concentrations varied by ±21% between technical replicate filters and by ±14% between duplicate assays. COVID-19 cases were correlated significantly (rho≥0.70) to wastewater SARS-CoV-2 RNA concentrations for seven plants, including large and small cities. SARS-CoV-2 data normalized to flow improved correlations to reported COVID-19 cases for some plants, but normalizing to a spiked recovery control (BCoV) or a fecal marker (PMMoV or HF183) generally reduced correlations. High frequency sampling demonstrated that a minimum of two samples collected per week was needed to maintain accuracy in trend analysis. We found a significantly different ratio of COVID-19 cases to SARS-CoV-2 loads in one of three large communities, suggesting a higher rate of undiagnosed cases. These data demonstrate that SARS-CoV-2 wastewater surveillance can provide a useful community-wide metric to assess the course of the COVID-19 pandemic.

## Introduction

The COVID-19 global pandemic has been challenging to control and marked by unchecked transmission and rapid spread. Community-wide burdens of COVID-19 have been difficult to assess, especially with limited availability of clinical testing at the beginning of the pandemic and spread by asymptomatic carriers [1,2], which has been reported to be as high as 30% [3,4]. These difficulties result in under-ascertainment of cases and complicate tracking the transmission progression through communities. Following the first reports of SARS-CoV-2 RNA detection in untreated sewage [5], wastewater surveillance has emerged as a promising tool for measuring community-wide levels of SARS-CoV-2, the virus responsible for COVID-19 [6]. As wastewater testing is not influenced by the availability of clinical testing resources, access to testing, or healthcare-seeking behavior, measuring SARS-CoV-2 concentrations in wastewater has the potential to offer insight into infection trends in the community that clinical testing might not capture.

Wastewater surveillance has the potential to offer cost-effective monitoring of SARS-CoV-2 prevalence at the population level, but there are still a number of challenges to overcome before it can be used effectively. Although SARS-CoV-2 is primarily a respiratory pathogen, it is also shed in feces at concentrations up to 10^7^ copies per gram [7,8], which allows the viral RNA to be detected readily in wastewater treatment plant influent. However, SARS-CoV-2 concentrations in feces vary widely among infected individuals and over the course of infection [7,8], which makes it difficult to extrapolate the number of infected individuals from wastewater concentrations. For example, using the reported shedding rates, which span five orders of magnitude, detection of the virus in wastewater could represent 100 to 6 million infected individuals [9]. Although empirical infection prevalence estimates have not yet been achieved with wastewater surveillance, retrospective analysis has demonstrated that SARS-CoV-2 RNA concentrations in wastewater treatment plant (WWTP) influent or wastewater solids correlate to clinical case trends [10–13]. Therefore, in instances when there are delays in reporting of diagnosed cases, wastewater can provide an early warning of later events such as hospitalizations or can indicate infection trends in communities with limited clinical testing resources [9,10,14,15].

One major challenge to implementing widespread SARS-CoV-2 wastewater surveillance is the lack of a standard method. Enveloped viruses partition between the solid and liquid phases in waste streams [16], and research is underway to evaluate concentration methods for these viruses in wastewater [17,18]. Using common controls can allow for comparison of results across different methods or laboratories [19]. A variety of surrogates have been used to estimate recovery, including bovine coronavirus (BCoV) [12,17], bovine respiratory syncytial virus (BRSV) [20], and human coronavirus (OC43) [21]. In addition, normalizing SARS-CoV-2 RNA concentrations to WWTP flow, the estimated amount of human fecal material in the samples (the fecal strength), and/or the population contributing to the viral load is needed to compare across samples and WWTPs. One promising candidate for normalizing SARS-CoV-2 concentration data is the pepper mild mottle virus (PMMoV), found in high concentrations in sewage [22]. This fecal marker is introduced into humans through the diet [23] and is attractive because, as a single stranded RNA virus, it may also serve as a RNA recovery control. Overall, it is critical to determine the technical variability in SARS-CoV-2 RNA measurements in day-to-day samples to enable timely interpretation of these data, i.e., How much of a change can be attributed to sampling or measurement variability versus changes in the community SARS-CoV-2 loads?

In this study, we examined the ability of wastewater surveillance to capture community trends in SARS-CoV-2 burdens. Intensive sampling was used to understand methodological variability and identify optimal sampling frequency that balances information gained with laboratory resources. We also assessed how recovery controls correlated to SARS-CoV-2 RNA detection and examined the usefulness of PMMoV and Human *Bacteroides* to normalize samples to fecal input. We also compared diagnosed COVID-19 cases to SARS-CoV-2 RNA concentrations in 12 WWTPs in Wisconsin, USA, from late summer 2020 to January 2021, which captured the November 2020 surge in cases reaching 100+ cases per 100,000 inhabitants, compared with ∼10 cases per 100,000 before the rise.

## Methods

### Sample collection

A total of 418 sewage influent samples were collected between August 30, 2020 and January 20, 2021 from 12 WWTPs in 10 cities in Wisconsin, United States (**Table 1**). Flow- or time-weighted composite samples were collected one to two times per week over a 24-hour period by each WWTP according to the plant standard collection procedures and then transferred to 250- or 500-mL bottles. Samples were stored at 4°C prior to overnight shipping on ice. Upon arrival at the laboratory, samples were stored at 4°C before processing within 5 h. Influent chemical/physical measurements were taken by WWTP operators employing their routine procedures.

**Table 1.** Description and details of operating conditions of the wastewater treatment plants (WWTP)

| WWTP | County | No. weekly samples | Population served | Capacity (MDG)<sup>1</sup> | Average daily flow (MDG, mean±SD)<sup>1</sup> | Composite frequency<sup>2</sup> (per MG) | TSS (mg/L, mean±SD) |
| --- | --- | --- | --- | --- | --- | --- | --- |
| Milwaukee Jones Island (JI) | Milwaukee | 2 or 5 | 470000 | 330 | 75±23 | 0.56 to 0.65 | 248±93 |
| Milwaukee South Shore (SS) | Milwaukee | 2 or 5 | 616000 | 300 | 74±17 | 1.70 | 282±116 |
| Racine (RAC) | Racine | 2 | 139000 | 36 | 15±4 | 0.08 | 127±31 |
| Waukesha (WAU) | Waukesha | 2 | 73000 | 39 | 7±1 | 0.10 | 505±303 |
| Brookfield (BRK) | Waukesha | 2 | 51000 | 13 | 8±1 | 0.35 | 211±48 |
| Green Bay (PLT) | Brown | 2 or 3 | 189000 | 49 | 24±4 | 0.09 | 230±97 |
| Green Bay De Pere (DP) | Brown | 1 or 2 | 43000 | 14 | 7±1 | 0.02 | 245±47 |
| South Milwaukee (CSM) | Milwaukee | 1 or 2 | 21000 | 6 | 3±1 | 0.25* | 196±84 |
| Port Washington (PTW) | Ozaukee | 1 | 12000 | 3 | 1±0 | 0.05 | 257±26 |
| Burlington (BUR) | Racine | 1 | 13000 | 4 | 3±0 | 0.07 | 296±51 |
| Cedarburg (CED) | Ozaukee | 1 | 12000 | 3 | 2±0 | 0.02 | 183±39 |
| Grafton (GRA) | Ozaukee | 1 | 11000 | 3 | 1±0 | 0.01 | 236±58 |

<sup>1</sup>Million gallons per day
<sup>2</sup>\*Frequency of sub-sample collection as sample pulse per million gallons (MG) or per hour for the City of South of Milwaukee.

We also conducted a three-week high frequency sampling campaign from September 13, 2020 and October 1, 2020 for the two Milwaukee WWTPs (JI & SS). For all three weeks, flow-weighted composite samples were collected daily from Sunday through Thursday (30 samples total; 5 days per week for 3 weeks at 2 plants).

### Sample filtration and nucleic acid extraction

Wastewater samples were mixed thoroughly by vigorous shaking, then 25 mL of each sample were pipetted into 50 mL tubes containing a magnesium chloride solution to reach a final concentration of 25 mM. We used BCoV that was obtained as the Calf Guard cattle vaccine (Zoetis, Parsippany, NJ) as a control for SARS-CoV-2 viral RNA recovery. The BCoV vaccine was rehydrated and 5 µL (about 100,000 cp/µL) was used for each sample filtration. See **Supplemental Text 1** and **Supplemental Figure S3** for additional details on the BCoV protocol and optimizations.

Samples were filtered in replicate for a subset of samples (167 of 418), including the high frequency sampling period. The 50 mL tubes were kept on ice until they were filtered through 0.8 µm cellulose esters HA filters (47 mm diameter; MF-Millipore, Carrigtwohill, Ireland). HA filters were transferred to a 2-mL ZR BashingBead Lysis tube (Zymo, Irvine, CA, USA) containing 650 µL of PM1 lysing buffer from the RNeasy PowerMicrobiome Kit (Qiagen, Hilden, Germany). BashingBead lysis tubes were placed at −80°C for a minimum of 2 h or up to overnight before nucleic acid extraction. While the filters were still frozen, 6.5 µL of β-mercaptoethanol (Sigma-Aldrich, St. Louis, MO, USA) was added to each filter tube. The BashingBead lysis tubes were then bead-beat using a mini beadbeater (Biospec Products, Bartlesville, OK, USA) in two runs of 2.5 min with a 5 min rest on ice in between and also after each run. After bead-beating, total nucleic acids (DNA and RNA) were extracted from the HA filters using the RNeasy PowerMicrobiome kit following the manufacturer’s instructions.

We also performed direct extractions (DE) on sewage influent samples and compared these to the samples processed using HA filters to assess HA filtering efficiency and recovery. Recovery of BCoV and PMMoV using HA filters was 7±4% and 45±26%, respectively (see **Supplemental Text 2**). The recovery efficiency of N1 and N2 could not be calculated owing to the unique sample above the limit of quantification from the direct extractions.

Optimization of filtration and extraction methods are described in **Supplemental Text 1**.

### Reverse transcriptase digital droplet PCR

We used one-step reverse transcription droplet digital PCR (RT-ddPCR) to target the N1 and N2 regions of the SARS-CoV-2 N gene using primer and probe sets designed by the Centers for Disease Control and Prevention (CDC) [24]. All reactions were set up following the one-step RT-ddPCR Advanced Kit for Probes (Bio-Rad, Hercules, CA) manufacturer’s instructions using the Mastercycler pro (Eppendorf, Hamburg, Germany) and the BioRad QX200 Droplet Digital System. Additional information is available in **Table S1**. Raw droplet amplification data were extracted from the QuantaSoft Analysis BioRad software and processed using the R package twoddpcr v1.11.0 [25]. For all assays, we ran a no template control using nuclease-free water. For the N1/N2 duplex assay, an additional positive control using a 1:8 diluted Exact Diagnostics *SARS-CoV-2* standard (Bio-Rad, Hercules, CA) was included in each run. The limit of detection (LOD) and limit of quantification (LOQ) were determined for the N1 and N2 assays (see **Supplemental text 1**). BCoV and PMMoV concentrations were determined in each sample using published assays [23,26]. Inhibition was evaluated in a subset of 75 samples by spiking in a known amount of BRSV into samples or DI water and quantifying by RT-ddPCR [20] (see **Supplemental Text 1** for details). No inhibition was observed for any samples tested.

### Human markers

Two human fecal markers, pepper mild mottle virus (PMMoV) [23] and human *Bacteroides* HF183 [27] were used to indicate the fecal strength of each sample (**Table S1**). The HB assay [28] was used to detect the HF183 marker. Sample extracts were diluted 1:10 for PMMoV and 1:100 for the HB assay. The PMMoV assay was performed according to the one-step RT-ddPCR procedure as described above; for each run, at least one no template control (NTC) well was added. Samples within the three-week high-frequency sampling period were run in duplicate for PMMoV and in single wells thereafter. The HB assay was run in duplicate for all samples using quantitative PCR (qPCR) as previously described [28]. For each HB assay run, positive controls, internal controls, and a NTC were used. The HB standard curve had a Y-intercept of 37.14, a slope of −3.37, and an efficiency of 99.36%.

### COVID-19 case data

COVID-19 cases for each WWTP service area were obtained by matching clinically confirmed COVID-19 case data (georeferenced) to WWTP sewershed boundaries in ArcGIS. Data was reported as cases per 100,000 people. The number of negative tests were also retrieved. The case rates (per 100,000) for the study sites are available through the Wisconsin Coronavirus Wastewater Monitoring Network (https://www.dhs.wisconsin.gov/covid-19/wastewater.htm).

### Statistical analysis

All statistical analysis was performed using R v3.6.3 [29]. Unless specified, we only used quantifiable data, i.e., above the LOQ, in analyses. To assess the importance of the assay, filter and consecutive day sampling on SARS-CoV-2 measured concentration variation, we calculated the relative standard deviation (RSD) between replicate assays, replicate filters, and from samples collected over two consecutive days, respectively. We evaluated the total measurement variability explained by the assay, filter, sample and WWTP factors using a nested variance component analysis from six WWTPs for which we evaluated replicate assays and filters. We conducted a one-way analysis of variance (ANOVA) followed by Tukey’s post-hoc test to explore the difference between BCoV recovery rate and PMMoV and HF183 concentrations. ANOVA was also used to assess the difference in the ratio of diagnosed cases per capita to SARS-CoV2 loads per capita. Monte Carlo simulations were used to evaluate the impacts of different sampling schemes on observed trends during the three-week high frequency sampling period at the Milwaukee JI facility. For the analysis below, N1 and N2 concentration below the LOQ were included. However, when all assays were below the LOD for a given sample, averaged concentrations were censored, i.e., LOD concentration was divided by two. Spearman’s rank correlation coefficient was used to determine the relationship between wastewater N1/N2 concentration and COVID-19 case data, as well as wastewater N1/N2 concentrations normalized by dividing to BCoV recovery rate, HF183 and PMMoV concentrations, or normalized per capita (N1N2×flow÷population served). To determine whether there is a lead time for wastewater concentration changes compared to clinical cases, we compared correlation coefficients between wastewater N1 concentrations and cases offset by −7 to +7 days. T-test was used to assess differences in correlations with a negative or positive offset. Statistical methodology is detailed in **Supplemental Text 3**.

## Results

### The SARS-CoV-2 N1 assay is more sensitive than N2

A significant positive correlation was observed between the N1 and N2 concentrations across all measured wastewater samples (Spearman’s rank correlation coefficient, rho = 0.86, n = 410, P < 0.001). The ratio of N1 to N2 in these samples was 1.83 ± 0.72 (mean ± SD). The shift from a single to double quenched probe for N2 improved the limit of detection (from 5 droplets to 3 droplets) due to a better discrimination between the negative and positive droplets (**Figure S1**). Even with this improvement, N2 was not as sensitive as N1, as the methodological change did not affect the limit of quantification (9 droplets for N2-single quenched probe while 10 droplets for the double quenched probe). Because of the high correlation between N1 and N2 and the efficiency of the N1 assay, we primarily focused on N1 concentrations for further analysis.

### Assessment of variability in SARS-CoV-2 detection and sources of variance for quantifiable data

To determine the sources and levels of variability in the N1 measurements, we first measured the

N1 concentration in assay replicates. Using only samples with concentrations above the LOQ, the median relative standard deviation (RSD) between N1 assay replicates was 14% (**Figure 1**). In duplicate filters the median RSD was 21%, while it was 23% for samples collected on two consecutive days (**Figure 1**).

**Figure 1.**
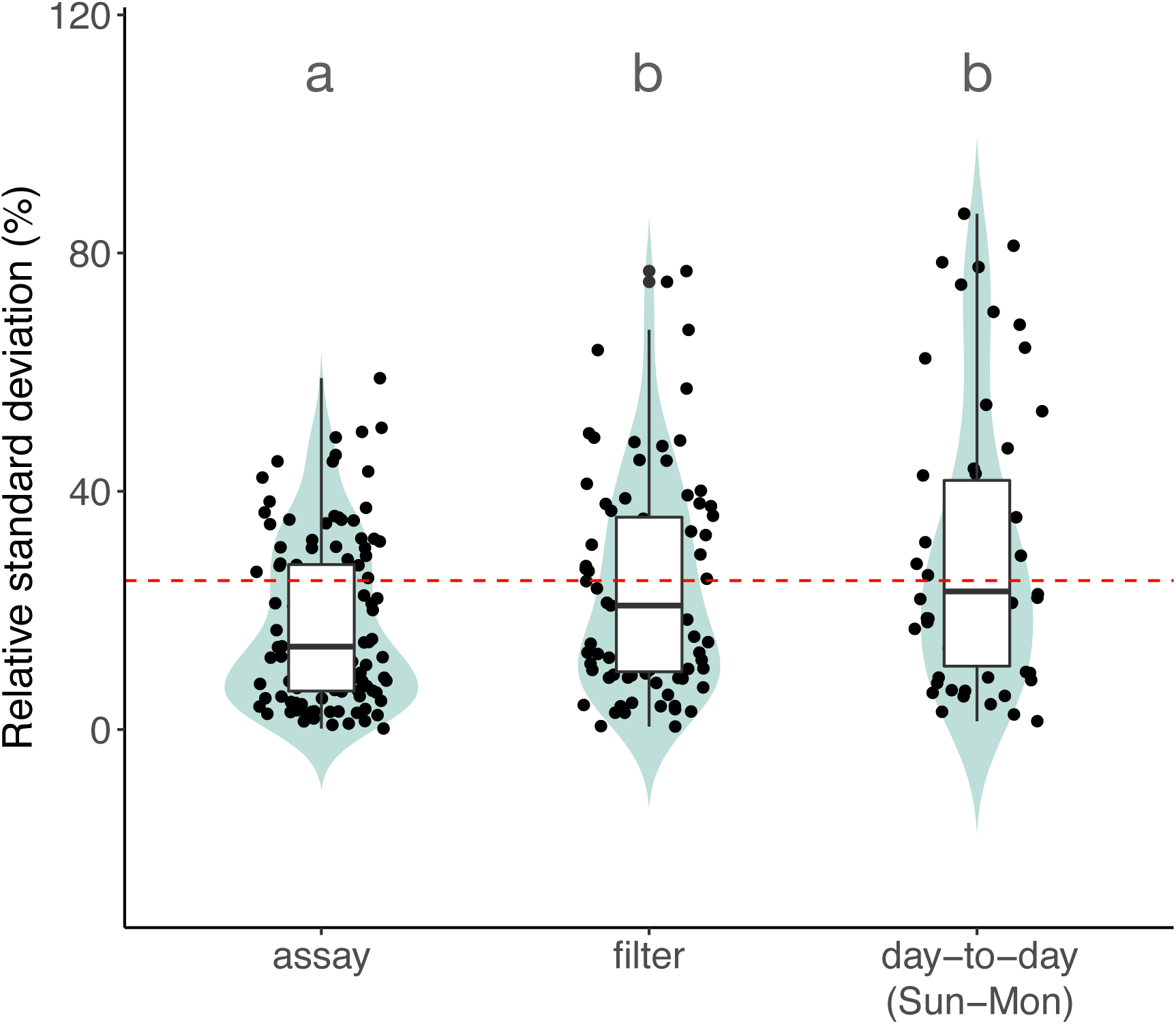
Distribution of N1 relative standard deviation percentages among assay duplicates (n=120 paired assay duplicates), filter replicates (n=83 samples in duplicate) and day-to-day collection (n=58 pairs of samples). The horizontal dashed line represents a 25% RSD. Only N1 concentrations above the LOQ were considered. Groups sharing the same letter are not significantly different (ANOVA, F(2) = 8.34, P < 0.001; Tukey-adjusted).

Nested variance component analysis was performed using quantifiable data to quantify and compare the variability in measured N1 concentrations introduced by the assay, filters, and between samples collected over two consecutive days (Sunday-Monday) (**Figure S2**). The total N1 variance was mostly attributable to the difference between samples (explaining 53% of the total variance), while the assay and filter factors contributed equally, explaining 24% and 22% of the total variance, respectively.

### Role of recovery controls in SARS-CoV-2 wastewater detection and quantification

The BCoV recovery rate across the time-series monitoring for all 12 WWTPs was on average 4.9 ± 4.2% (mean ± SD). However, a significant difference was observed between the facilities (**Figure S4**, ANOVA, F(11) = 26.91, P < 0.001), with some WWTPs having more consistent recovery than others: e.g., Waukesha ranged from 0.21 to 3.0% (n=41) compared with Milwaukee Jones Island, which ranged from 0.89 to 28.0% (n=51). This recovery was not correlated to the total suspended solid concentrations in samples (Spearman, n=381, rho = −0.25).

We also examined the recovery of PMMoV from WWTP samples. The concentration across all samples was on average 7.2 ± 0.3 log_10_ cp/L (geometric mean ± SD). Similar to BCoV, a significant difference in PMMoV concentrations was observed between the WWTPs (**Figure S4**, ANOVA, F(11) = 10.76, P < 0.001). Direct extraction of wastewater samples also resulted in similar differences in PMMoV concentrations between WWTPs (see **Supplemental Text 2**), suggesting that these differences represent variation in human fecal marker loads and were not a function of methodological variability.

Samples processed on duplicate filters were used to assess the power of the recovery controls to correct for within-sample SARS-CoV-2 concentration variation. For this analysis, we compared the RSD of BCoV recovery rate and PMMoV concentrations to N1 concentrations between replicate filters of the same sample. Both BCoV and PMMoV poorly explained N1 variation (Spearman’s rank correlation coefficients, BCoV, n = 62, rho = 0.11; PMMoV, n = 55, rho = 0.17), suggesting neither of these constituents mimics recovery of the SARS-CoV-2 enveloped virus.

### High frequency sampling to determine optimal sampling design

During our high-frequency data collection (i.e., 5 days per week with replicate filtration), we observed a steady and significant increase (∼0.5 log_10_ cp/100 mL) in N1 concentrations at the Jones Island facility that corresponded to an increase in clinical case levels, allowing us to investigate the impact that a reduced sampling frequency and altered sampling scheme (days of the week) had on this observed trend (see **Supplemental Text 3**). We created a series of Monte Carlo simulations where, in each case, data points were simulated using a uniform distribution between the minimum and maximum SARS-CoV-2 N1 concentrations observed on each day and reducing either the number of replicates or the number of days sampled. By reducing to single data points per day (i.e., simulating a single filtration as opposed to duplicate filtration), a significant trend was still observed in >99% of simulations.

The number of days sampled were adjusted either by clustering sampling at the beginning of the week or by spreading sampling throughout the week at different frequencies. By clustering samples at the beginning of the week, a reduction to 4 or 3 days of sampling (compared to 5) had negligible impact on the observed trend, with >99% of simulations still resulting in a significantly increasing concentration (**Figure 2**). However, sampling only on one or two days weekly at the beginning of each week resulted in a greater loss in significance for the observed trends simulated, with 62% and 44% of simulations resulting in no observable increased trend for one or two days sampled, respectively. By spreading sampling dates through the course of a week, a Sunday/Tuesday/Thursday sampling scheme resulted in no impact on the observed trend, while a Monday/Wednesday sampling scheme resulted in a loss of the observed trend in 14% of simulations.

**Figure 2.**
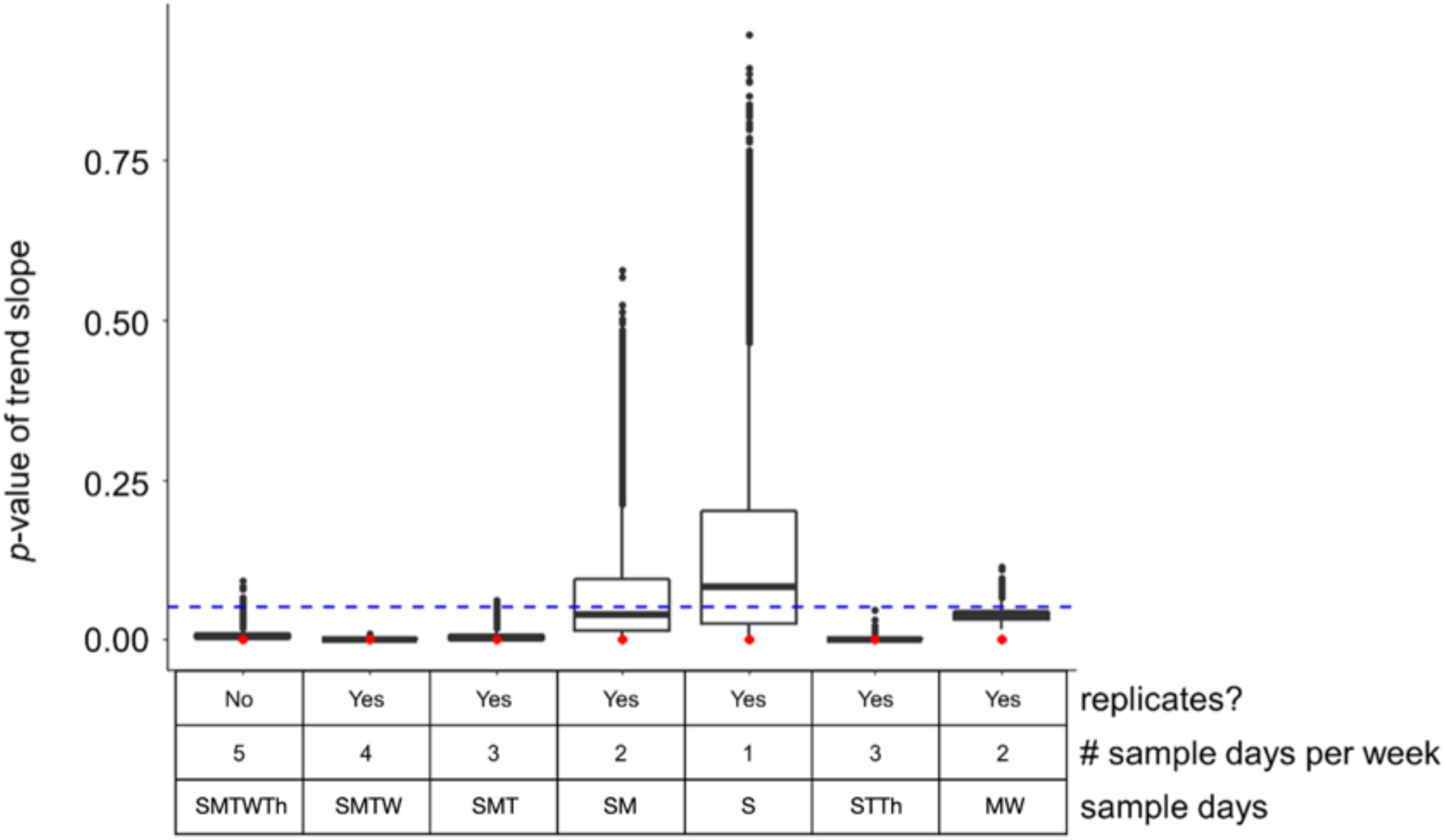
Significance (*p*-values) of trendline slopes from Monte Carlo simulations. Each bar represents 10,000 simulations of the described sampling scheme (x axis). Red dots represent the slope *p*-value using all original data collected, and the blue dashed line marks a significance level of 0.05.

### Fluctuation in levels of bacterial and RNA viral human markers

The HF183 human marker concentration was on average of 8.8 ± 0.2 log_10_ cp/L (geometric mean ± SD) across all WWTPs, which was overall about an order of magnitude higher than PMMoV (**Figure 3**). Both human fecal markers exhibited relatively consistent concentrations over time in a single WWTP (within an order of magnitude variation). However, HF183 concentrations were less variable than PMMoV across all WWTPs (HF183 RSD = 53% vs. PMMoV RSD = 184% for HA filters or 102% for direct extraction). Overall, neither of the human markers correlated to the flow for each plant (**Figure S5**). No trend was observed between decreasing variability of either marker or increasing population size, suggesting that the variability was not driven by sample size of the population.

**Figure 3.**
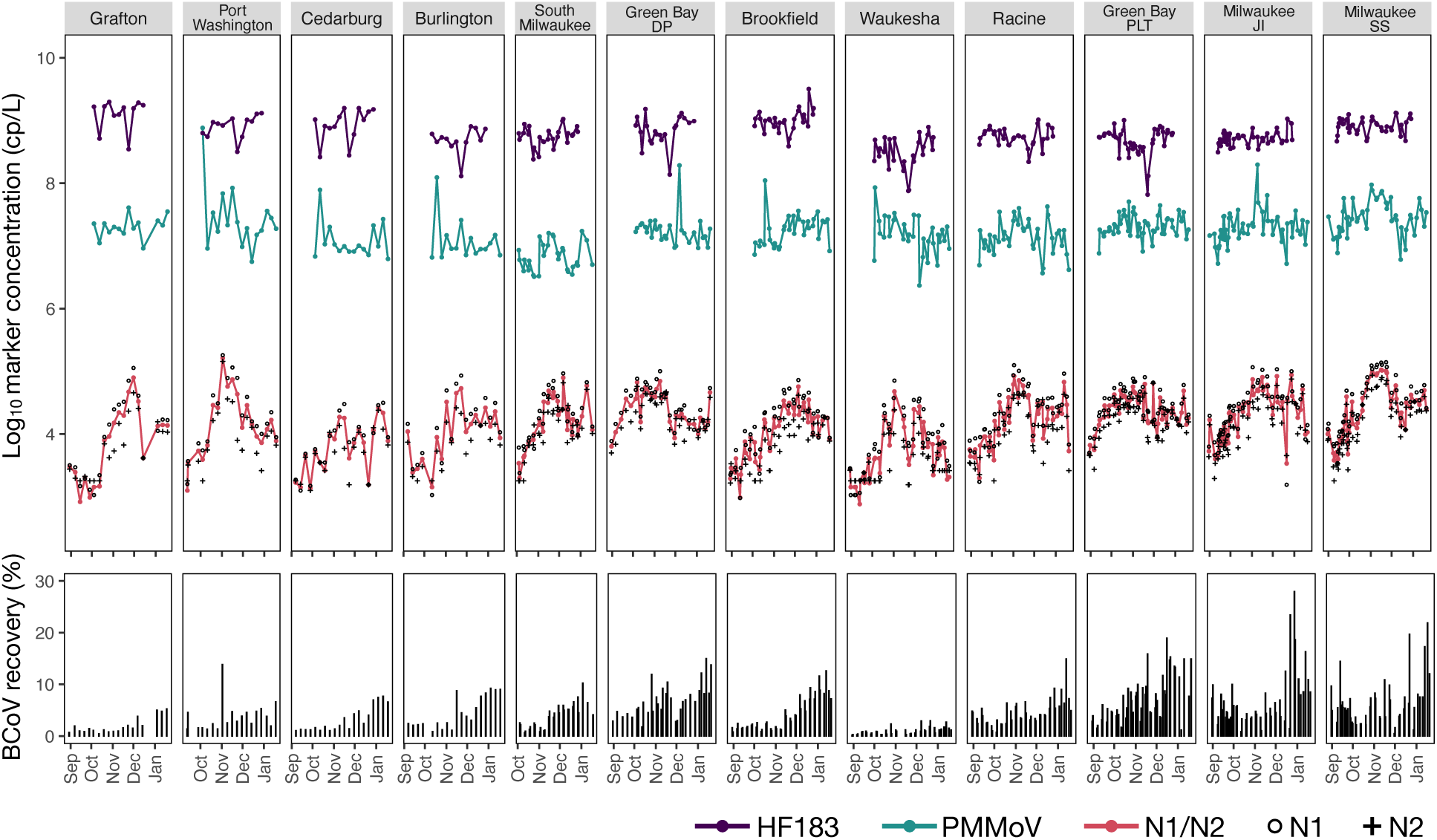
Time-series of censored N1/N2, PMMoV and HF183 average concentrations (upper panel) and BCoV recovery rate (lower panel) across 12 WWTPs. WWTPs are sorted from the smallest (Grafton) to the largest (Milwaukee South Shore) population size served.

### Correlation of SARS-CoV-2 concentrations to COVID-19 case rates and effects of normalization to human fecal markers, flow, or recovery

Using aggregated address level clinical case reports (by date of test), we determined the correlation of censored SARS-CoV-2 wastewater concentrations with the 7-day moving average of diagnosed cases of COVID-19 (**Figure 4**). Spearman rank correlations ranged from rho=0.58 to 0.87, with no relation to the size of the WWTP.

**Figure 4.**
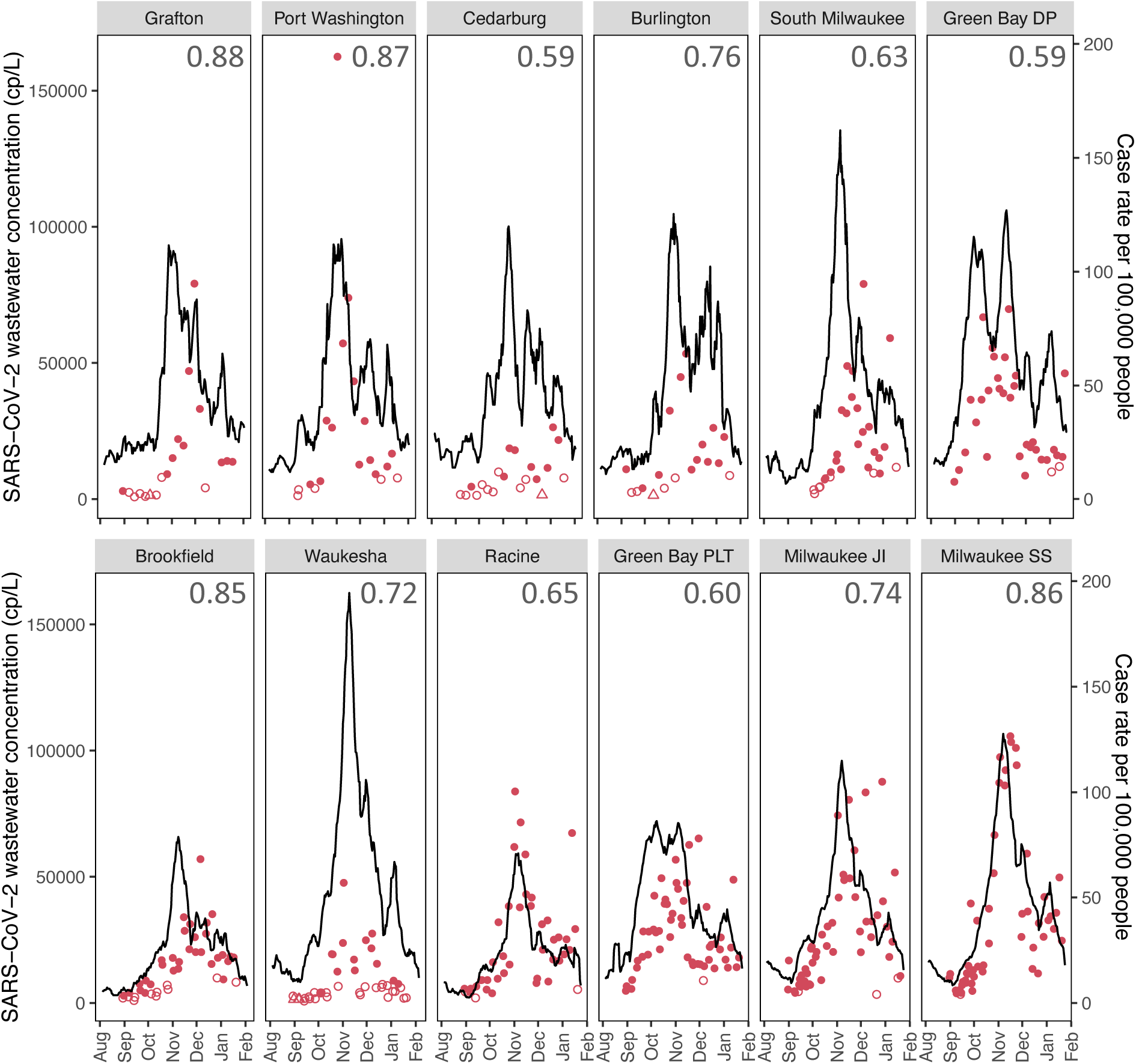
Time-series of censored N1/N2 averaged per day and the 7-day moving average of COVID-19 cases per 100,000. Wastewater treatment plants are ordered from largest to smallest based on population served. Values display the Spearman’s rank correlation coefficients between SARS-CoV-2 wastewater concentrations and case rate per capita. Open circles and triangles indicate BLQ and BLD concentrations, respectively. WWTPs are sorted from lowest (Grafton) to highest population served (Milwaukee South Shore).

We also examined correlations of SARS-CoV-2 wastewater measurements to COVID-19 cases after normalizing to either flow or the human fecal markers, PMMoV or HF183 (**Figure 5**). Correlations were only improved slightly when adjusting to flow and only in select cities. For example, the average correlation across the 12 WWTPs was rho=0.73 based on raw concentrations and 0.74 when adjusted to flow. Correcting for BCoV recovery, HF183, or PMMoV actually decreased the correlation coefficient in many cases, with average correlations falling to 0.59, 0.66 and 0.56, respectively. Three-day moving averages were also examined and did not increase the strength of the correlation (**Figure S6**).

**Figure 5.**
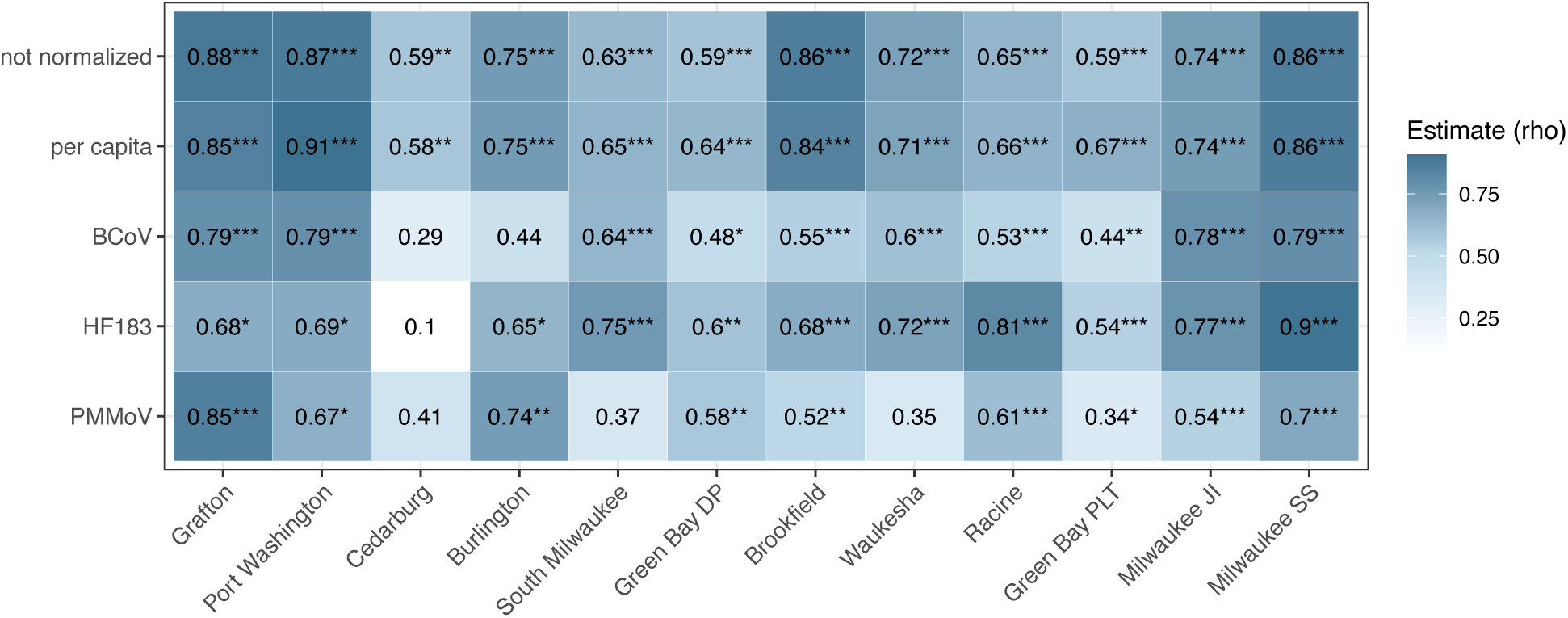
Correlations between 7-day moving average of diagnosed COVID-19 cases and unadjusted average N1N2 targets for SARS-CoV-2 wastewater concentrations, and concentrations normalized to gene copy per capita (concentrations x flow)/sewershed population, BCoV recovery, HF183 or PMMoV concentrations. WWTPs are ordered from smallest to largest population served. *P-value* significance level: *0.05, **0.01, ***0.001.

Our correlations used the date of the clinical test compared with wastewater concretions on that date, which removes effects of reporting delays on the correlations. Using flow normalized data, we examined whether correlations improved with cases moved to an earlier date (−1 to −7) or to a later date (+1 to +7 days) for four of the largest WWTPs. This analysis essentially moves the clinical case curve forward or backward from the SARS-CoV-2 RNA concentrations. We found that the correlations slightly improved as the cases were offset to a later date, but optimal timeframes were different for each WWTP (0 to +6 days; **Figure S7**). The correlations were significantly lower for the −1 to −7 offset compared with the +1 to +7 day offset (t-test, P<0.05).

### Wastewater SARS-CoV-2 concentrations indicate undiagnosed cases

Using case data from the sewershed boundaries of three largest communities, we examined whether the ratio of identified cases per 100,000 residents to wastewater SARS-CoV-2 concentrations (million gene copies (MGC)/person per day) was consistent across the WWTPs. Two of the communities are serviced by two WWTPs, which provides two different measurements within the community. We hypothesized that a comparatively lower cases:wastewater concentration ratio would indicate a higher number of undiagnosed cases. We found that Green Bay DP had the highest ratio, which was not significantly different from Green Bay PTL, Milwaukee SS, and Milwaukee JI (**Table 2**). Conversely, Racine had a significantly lower ratio compared to these other three WWTPs (**Table 2**, P ≤ 0.031). Testing coverage mirrored this trend as Green Bay DP had the highest number of tests per person and Racine had the lowest, supporting that the low ratio observed in Racine could be associated with undiagnosed cases.

**Table 2.** Comparison of diagnosed cases per capita with SARS-CoV-2 RNA per capita across the large communities.

| WWTP | Cases in Sewershed<sup>1</sup><br>(per 100,000 people) | N1 copies<sup>1</sup><br>(MGC/person/day) | Average of daily ratios<br>Cases:N1 copies | Testing coverage<br>Clinical tests/person |
| --- | --- | --- | --- | --- |
| Green Bay DP | 84 | 21 | 4.96 | 0.90 |
| Green Bay PLT | 56 | 17 | 3.81 | 0.61 |
| Milwaukee JI | 55 | 19 | 3.63 | 0.65 |
| Milwaukee SS | 57 | 18 | 3.97 | 0.65 |
| Racine RAC | 47 | 23 | 2.83* | 0.56 |

<sup>1</sup>Average of study time period August 30, 2020 to January 20, 2021
\*Significantly different compared to the other four WWTPs (ANOVA, F(4) = 6.467, P<0.001; Tukey-adjusted, P ≤ 0.031).

## Discussion

In this study, low technical variability and strong correlation to case reports show the utility of SARS-CoV-2 wastewater concentration data for providing a biologically independent measure of SARS-CoV-2 infections across a community. Our results indicate the HA filter method [30,31] combined with one-step RT-ddPCR is effective for tracking SARS-CoV-2 concentrations in wastewater influent. Similarly to reports by LaTurner et al [17], we found the HA filter method was relatively fast and allowed an effective analysis volume for each ddPCR reaction (∼1.5 mL), which provided the needed sensitivity to quantify SARS-CoV-2 when reported cases were as low as 9 per 100,000 people in the watershed. We also found that the N1 assay was more sensitive than the N2 assay when levels were near the limits of detection, likely because of assay efficiency. Further, the ddPCR method is reported to be more sensitive than qPCR [12], and combining multiple wells can increase the effective sample volume analyzed. As community transmission decreases, more sensitive measures might be needed since early in the pandemic SARS-CoV-2 RNA levels were near or below the limits of detection [13,30]. Analysis of primary solids might be useful for detection as recent reports show ∼300-3000 fold increase in sensitivity [12]. Methods to detect and quantify SARS-CoV-2 RNA in the complex matrix of wastewater are time consuming and expensive, and balancing the need for technical replicates to define confidence in measurements with limited resources and the need for timely data is a reality for many laboratories. In a large comparison study of 31 laboratories, technical variability within a given standard operating procedure (SOP) for measuring wastewater SARS-CoV-2 was below ±1.15 log value for 80% of the SOPs [6], so measurement inconsistency can be fairly high [8], but this comparison study was conducted early in the pandemic when many laboratories were still optimizing and testing multiple methods. Here, we examined the variation in a consistent method, testing nearly 40% of our 418 samples in duplicate over nearly six months. We found the overall variability between replicate filters from the same sample to be relatively low (0.3 log value), and RT-ddPCR assays contributed an equivalent amount of variability as replicate filters from the same sample. Therefore, we suggest that once methods are evaluated and found to be acceptable, replicate filters are not necessary for reliable tracking of wastewater SARS-CoV-2 concentrations.

Our methods assessment also revealed 24-hour composite samples provide representative measurements of SARS-CoV-2 RNA concentrations. We assumed that SARS-CoV-2 burdens in the community change minimally from one day to the next and examined samples collected two days in a row. We found good agreement between samples. This is remarkable since flows into WWTPS are millions of gallons per day, and composite samples collect 1-2 L, with 25 mL analyzed on a single filter. WWTP influent is assumed to be a well-mixed sample of contributions from the human population and previous studies in our laboratory [32,33] have demonstrated that human fecal markers are at steady-state levels in WWTP influent, but for these markers, nearly every human in the service area is contributing. In the case of SARS-CoV-2 infections, only a small number per 100,000 people are contributing SARS-CoV-2 based on clinically diagnosed cases, albeit these data likely underestimate prevalence [34]. We note that our analysis of consecutive days examined primarily large sewersheds servicing more than 50,000 people. While the two smaller WWTPs also had good agreement in samples collected two days in a row, further analyses in smaller sewersheds are needed to understand the effect of size on sample representativeness.

Identifying a sampling frequency for wastewater surveillance data that captures case dynamics is another important aspect of maximizing limited resources while making wastewater data meaningful and actionable for public health entities. We found that the least resource-intensive sampling scheme that maintained a high degree of confidence in observing moderate concentration changes (∼0.5 log_10_ copies/100 mL) was two non-consecutive days per week. Our results also indicate that sampling frequency is more critical for data interpretation than analysis of replicate filters. We recognize that our analysis is limited by the dynamics specific to the three-week period analyzed and the service area, but our findings agree with those by Graham et al. [12], who assessed wastewater solids and also concluded that sampling two days a week captured case trends equivalent to every day sampling.

Wastewater is complex and can be very different in composition across WWTPs. Proper controls are needed to verify the quality of the measurements and to compare between methods or WWTPs. Similar to Graham et al. [12], we found that BCoV did not mimic the behavior of the SARS-CoV-2 virus in our replicate filters. We also found that normalizing SARS-CoV-2 RNA concentrations to recovery decreased correlations to case data (**Figure 5**). This might be expected since enveloped viruses like SARS-CoV-2 partition into solids in wastewater [16] and would have more contact time than a spiked control. In this study, we found lengthening the contact time of BCoV with wastewater from 30 minutes to a maximum of 48 hrs did not yield improved BCoV recovery (**Figure S3**), suggesting additional factors specific to the sewer environment may be influencing recovery differences between SARS-CoV-2 and surrogate viruses used as recovery controls. We also found that in general, samples with higher TSS did not yield higher SARS-CoV-2 concentrations. While BCoV recovery did not correlate to N1 or N2 recoveries, we did find that a very low BCoV recovery (e.g., <1%) was a reliable flag for method failure and typically resulted in very low or absent N1 or N2 detection.

We found that normalizing to waste stream characteristics (flow, human markers) had little effect on correlations to case data (**Figure 5**). Normalizing to flow slightly increased correlations, but not for every plant. Adjusting to the amount of fecal contribution coming into the WWTP using PMMoV or HF183 actually decreased correlations in most cases, which might suggest that variability across measurements (SARS-CoV-2, fecal markers) is a larger influence than differences in the fecal loads of samples. These fecal markers are likely more useful in instances were fecal loads would be highly variable, such as specialized sampling within the sewershed. PMMoV is attractive as a normalizing factor since it can be used simultaneously as an RNA recovery control. However, as the expected population sampled decreases, the variability in this diet-driven marker needs to be considered since individual contributors would more strongly influence measured concentrations [35].

SARS-CoV-2 wastewater concentrations strongly correlated to diagnosed case rates, similar to other reports [10–12,35]. Sewershed boundaries were used by the Department of Health Services to assign caseloads and create de-identified data, which increased the precision of these analyses. Similar to reports by Graham et al. [12], we found trends mirrored clinical cases in all 12 WWTPs across both increasing and decreasing clinical case trends. We also found wastewater was not a leading indicator when the date of the test was used, suggesting that infected individuals did not contribute SARS-CoV-2 at appreciable levels to wastewater at appreciable levels before diagnosis. However, case reporting can take several days or even weeks for that information to reach public health officials, and in such cases, wastewater could provide more timely information [14]. When case rates (by date of the test) were lagged in a positive direction (forward in date), correlations slightly improved, suggesting that diagnosed cases contribute for several days to the SARS-CoV-2 burdens. We also hypothesized that comparing the per capita case rate with SARS-CoV-2 loads in wastewater could be used as an undiagnosed cases indicator. We did find that the one community with significantly lower ratios of diagnosed cases:SARS-CoV-2 loads also had a low testing rate, which would support this hypothesis. Understanding these relationships will be useful when clinical testing wanes; however, more work is needed to validate these comparisons across time within and across communities.

## Conclusions

SARS-CoV-2 wastewater surveillance can be highly effective for estimating changes in community COVID-19 burdens. We found that our sampling scheme of two days captured trends and our validated method had low variability, which improves confidence in interpreting SARS-CoV-2 RNA concentrations. For this new tool to be useful to public health practitioners, key information should accompany data, including information on recovery controls, inhibition, and the technical variability in the method [19,36]. We were also able to establish the degree to which diagnosed cases correlated to the SARS-CoV-2 wastewater data, which varied across communities and should be considered when interpreting possible trends. While the pandemic continues to evolve, sensitive methods will be crucial, and may include concurrent strain surveillance [37,38] and incorporation of models to estimate transmission dynamics [39].

## Data Availability

Data is available in Supplemental Dataset1 and at https://www.dhs.wisconsin.gov/covid-19/wastewater.htm

## Acknowledgments

We thank the Office of Health Informatics and the Bureau of Information Technology Services at the Wisconsin Department of Health Services for their contributions mapping COVID-19 case data and making information accessible to the public through the DHS COVID-19 Wastewater Monitoring website. We thank Elexius Passante and Melissa Schussman from the McLellan lab and Angela Schmoldt from the Great Lakes Genomics Center for contributing to method evaluation and data generation. We thank Jeanne Li, UW-Madison Data Sciences Program, for statistical consulting. We also thank the participating wastewater treatment plants for providing samples and sharing expertise on individual systems. The project was supported with funding from the Wisconsin Department of Health Services through the CARES Act and through Student Fellowship funds from the Milwaukee Metropolitan Sewerage District.

