## Supplemental Information for "Evaluation of sampling frequency and normalization of SARS-CoV-2 wastewater concentrations for capturing COVID-19 burdens in the community"

Number of pages: 14
Number of tables: 2
Number of figures: 7

**List of Supplemental Text, Tables, and Figures**

**Supplemental Text 1** Materials and Methods supplements and optimizations

**Supplemental Text 2** Comparison of direct extraction recoveries with HA filter recoveries

**Supplemental Text 3** Details for statistical methods

**Table S1** List of the ddPCR and qPCR assays

**Table S2** Influence of the incubation time on BCoV recovery rates and N1 and N2 concentrations

**Figure S1** Two-dimensional RT-ddPCR cluster plots of N1/N2 duplex assays using single- and double- quenched N2 probes

**Figure S2** Nested variance component analysis results for assays, filters, samples, and WWTP

**Figure S3** Influence of BCoV incubation time (with or without shaking) on N1/N2 detection and BCoV recovery

**Figure S4** Distribution of (a) BCoV recovery rates and (b) PMMoV concentrations averaged per day and per WWTP

**Figure S5** Relationship between the concentrations of human markers (a) PMMoV and (b) HF183 and the average daily flow at the WWTPs

**Figure S6** Spearman's rank correlation coefficients between case rate per 100,000 people averaged per 7-day or 3-day and the N1 or N2 concentration in sewage without and without normalization

**Figure S7** Spearman's rank correlations of case date (by the date of test) and SARS-CoV-2 million gene copies (MGC) per person in sewersheds

**Supplemental Text 1: Materials and Methods supplements and optimizations**

Filtration and extraction protocols are available on protocols.io: dx.doi.org/10.17504/protocols.io.bpg7mjzn and dx.doi.org/10.17504/protocols.io.bpg5mjy6, respectively.

***Limits of blank, detection, and quantification*** For the N1 and N2 assays, we defined the limit of detection (LOD) as the lowest analyte concentration likely to be reliably distinguished from the limit of blank (LOB) and at which the detection is feasible, according to (Armbruster and Pry 2008). The LOB was determined by quantifying N1 and N2 in a pre-pandemic sewage influent sample collected in August 2019. The LOB was calculated as the maximum value from a 90% confidence interval of the average false positive droplets across 24 replicate measurements. If the LOB was zero (i.e., no positive droplet in any of the replicates), then the LOD was set at 3 positive droplets as a conservative threshold. Serial dilutions of the Exact Diagnostics SARS-CoV-2 standards (including 200 cp/µL of the N gene; Bio-Rad, Hercules, CA) were run in triplicates to determine the limit of quantification (LOQ). We define the LOQ as the lowest concentration at which the analyte can not only be reliably detected but also some predefined goals for bias and imprecision are met (Armbruster and Pry 2008). For N1 and N2 we set the LOQ as the concentration at which the relative standard deviation was ≤30% between triplicate assay measures and the difference between the calculated and expected concentrations was ≤30%. The LOB, LOD, and LOQ were always rounded to the nearest higher integer. For both the one-step RT-ddPCR N1 and N2 assays, the LOD was set at 3 positive droplets (LOB was 0) and the LOQ was 10 positive droplets. This was applied to both N1 and N2 assays except for the N2 assay run with the single quenched probe (BHQ1), for which the LOB was 4 positive droplets, the LOD was 5, and the LOQ was 9.

***Bovine coronavirus (BCoV) internal recovery control*** Bovine coronavirus solution was prepared out of one dose of freeze-dried Calf Guard cattle vaccine (Zoetis, Parsippany, NJ) rehydrated in 3 mL of sterile 1X TE buffer and aliquoted into 100 µL stock solution aliquots, which were stored immediately at -80 °C. To use, the BCoV stock solution aliquot was thawed on ice and vortexed thoroughly; each aliquot was used for a maximum of two freeze-thaw cycles. A volume of 5 µL of the BCoV stock solution (about 100,000 cp/µL, titer determined by dilution series) was spiked in to the sample before filtering. To define the concentration of BCoV stock solution, 5 µL were extracted using the RNeasy PowerMicrobiome kit as described for direct extraction (see Supplemental Text 2), except that the BCoV stock solution was transferred into 2-mL tubes containing 500 µL of warmed PM1 solution, 5 µL of β-mercaptoethanol and 150 µL of IRS solution. The extracted BCoV RNA was then serially diluted 1:2 for four dilutions and run in duplicate using the BCoV assay [1] according to the one-step RT-ddPCR procedure as described in the main text. In samples, the BCoV recovery was calculated as the ratio of the final quantity measured to the quantity added to each wastewater sample and was used to estimate the loss of the RNA during sample processing. A detailed protocol of the BCoV solution preparation and the titer estimation is available at protocols.io: dx.doi.org/10.17504/protocols.io.bpg8mjzw.

We observed that BCoV recoveries did not correlate to N1 or N2 recoveries when comparing replicate filters (Spearman's rank correlation coefficients, BCoV, n = 62, rho = 0.113, see main text). Because BCoV is an artificial spike-in control, we examined if the time from spiking into a sample to processing that sample impacted its recovery since enveloped viruses are expected to adsorb onto solids in the wastewater matrix. We hypothesized that the lack of congruence between BCoV recoveries and N1 or N2 recoveries was a result of the longer time that SARS-CoV-2 interacted with the wastewater matrix. We found that different incubation times and agitation regimes had no effect on N1 and N2 recoveries (Figure S3). However, no incubation at all generated slightly better BCoV recoveries. Results were confirmed across samples from several wastewater treatment plants (WWTPs) (Table S2).

***Inhibition control Bovine Respiratory Syncytial Virus (BRSV)*** Bovine Respiratory Syncytial Virus (BRSV**)** solution was prepared from 25 doses of freeze-dried Inforce 3 Cattle Vaccine (Zoetis, Kalamazoo, MI, USA) rehydrated in 5 mL of sterile 1X TE buffer and aliquoted into multiple 150 µL stock solution aliquots, which were stored immediately at -80 °C. A total of 150 µL were extracted using the RNeasy PowerMicrobiome kit as described for the direct extraction (see Supplemental Text 2). The extracted BRSV RNA was then serially diluted 1:10 in water for four dilutions and run in duplicate using the BRSV assay [2] according to the one-step RT-ddPCR procedure as described in the main text. Approximately 4000 copies of BRSV were spiked into each RT-ddPCR reaction to evaluate the potential inhibition of PCR amplification. BRSV was also spiked into two wells containing only sterile PCR grade water to serve as a no inhibition reference measure. No template control wells using PCR grade water were also included. Reaction inhibition in the wastewater samples was identified as the ratio of BRSV concentrations in the reference wells to the wastewater samples. If the BRSV concentration in the wastewater samples was < 50% of the reference wells, then the reaction was considered to be inhibited. No inhibition was observed among the 75 wastewater samples tested.

***Filter freezing duration prior to extraction*** We examined the effect that freezing HA filters at -80 °C prior to extraction had on virus recovery. To measure this, we split 24-hr composite wastewater samples into two subsamples, one that was filtered and extracted as soon as it was transferred to the lab and another that was filtered and then kept overnight at -80 °C prior to extraction. No significant difference was observed for the SARS-CoV-2 concentrations (N1 assay) or BCoV recoveries between the subsamples (N1: paired Wilcoxon-test, $V = 9$, $P = 0.844$; N1: paired t-test, $t = -0.446$, $P = 0.674$; $n = 6$; BCoV: paired t-test, $t = -1.483$, $P = 0.198$).

### Supplemental Text 2: Comparison of direct extraction recoveries with HA filter recoveries

***Direct extraction protocol*** For some wastewater influent samples, results from virus concentration methods were compared to those from a direct extraction (i.e., no concentration step). The RNeasy PowerMicrobiome Kit (Qiagen, Hilden, Germany) was used to extract RNA for all direct extraction samples. To begin, 125 - 150 µL of wastewater was transferred into a 2-mL tube containing 400 µL of warmed PM1 solution. Four microliters of β-mercaptoethanol (Sigma-Aldrich, St. Louis, MO, USA) was then added to each tube. The mixture was vortexed 15 seconds, and then incubated at room temperature for 10 min. The manufacturer's instructions were then followed, except that 100 µL of solution IRS was used. Sample RNA was eluted in 60 µL of RNase-free water.

To assess the efficiency of our HA filtering method, we compared the recoveries of BCoV and pepper mild mottle virus (PMMoV) in samples that were processed with both HA filters and direct extraction. On average, the recoveries were correlated, but with lower recovery efficiency for BCoV on HA filters. The ratio of HA copies to direct extraction copies for BCoV was 7±4% (Spearman, $n = 106$, rho = 0.003) and 45±26% for PMMoV (Spearman, $n = 106$, rho = 0.383). The recovery efficiency of N1 or N2 could not be calculated owing to the low number of samples ($n = 1$) above the limit of quantification from the direct extraction.

### Supplemental Text 3. Details for statistical analysis

Unless specified, we only used quantifiable data, i.e., above the LOQ in analyses. For samples processed with replicate filters and/or replicate assays from the same filter, concentrations were first averaged by assay replicates per filter and then by filters per sample. Concentrations were $\log_{10}$-transformed prior to analysis as indicated.

A comparison between the N1 and N2 assay concentrations was performed only on samples where both assay concentrations were above the LOQ. To assess the importance of the (i) assay, (ii) filter, and (iii) consecutive day sampling on SARS-CoV-2 variability detection, we calculated the relative standard deviation (RSD), i.e., the ratio of the standard deviation to the mean between replicate assays (from the same filter), duplicate filters (from the same sample), and from samples collected over two consecutive days (from the same WWTP), respectively.

We focused on Sunday and Monday samples because they represented 60% of our total day-to-day samples.

A nested variance components analysis was performed on $\log_{10}$-transformed data from six WWTPs, where we had consecutive day samples processed in duplicate filters with each filter quantified in replicate assays, to evaluate the total variance explained by the assay, filter, sample and WWTP factors on the SARS-CoV-2 variability. The analysis was done using the function *anovaVCA* implemented in the R package VCA v1.4.3 [3]. A nested analysis of variance (nested ANOVA) was also performed on these data using the function *aov* to examine the significance of individual factors in explaining the observed variability, as well as to check the normality of residuals (Figure S2). One-way ANOVA followed by Tukey's post-hoc test was performed using the function *aov* to explore the difference of BCoV recovery rate and PMMoV and HF183 concentrations across the WWTPs. The normality of residuals was evaluated for each analysis. We used Spearman's rank correlation coefficient to measure the degree of correlation between qualitative variables.

Monte Carlo simulations were used to evaluate the impacts of different sampling schemes on observed trends during the 3-week high frequency sampling period at the Milwaukee JI facility. For these simulations, each day's SARS-CoV-2 N1 concentrations were represented by a uniform distribution between the minimum and maximum N1 concentrations detected across all filter and assay replicates for that date. For these simulations, each day's SARS-CoV-2 N1 concentrations were represented by a uniform distribution between the minimum and maximum N1 concentrations detected across all filter and assay replicates for that date. Simulated datasets were then constructed by randomly selecting values from these distributions based on seven different sampling schemes: (1) SMTWTh but no replicates, (2) SMTW, (3) SMT, (4) SM, (5), S, (6) STTh, and (7) MW. Sampling schemes 2-5 cluster sampling dates near the beginning of the week, which is often logistically favorable to maximize weekday sample processing time after sample collection. For each simulation, values were randomly selected from the uniform distributions representing the appropriate sampling dates, a linear trend was fit to the data, and fit parameters were extracted. This was repeated 10,000 times for each of the seven sampling schemes. A linear trend was considered to be observed if the *p*-value of the slope was $< 0.05$.

One-way ANOVA analysis was used to assess the significance of differences between BCoV recovery rates among WWTPS, as well as the ratios of diagnosed case rates per 100,000 to SARS-CoV-2 million gene copies (MGC) per person. Normality and homoscedasticity of ANOVA model residuals were evaluated using normal QQ plots and residual plots. If needed, dependent variables were $\log_{10}$-transformed prior to analysis.

Spearman rank correlations were used to determine the correlations of wastewater SARS-CoV-2 concentrations and the concentrations normalized to BCoV, WWTP flow, or PMMoV. To determine whether there is a lead time for wastewater concentration changes compared to clinical case changes we examined correlation coefficients between wastewater N1 concentrations and cases shifted forward 7 days or backward 7 days.

**Table S1** List of the RT-ddPCR and qPCR assays

| Assay | Sequence 5'-3' | Suppliers | PCR | Annealing temperature | Reference |
|---|---|---|---|---|---|
| N1 | F: GACCCCAAAATCAGCGAAAT<br>R: TCTGGTTACTGCCAGTTGAATCTG<br>P: FAM/ACCCCGCAT/ZEN/TACGTTTGGTGGACC/IABkFQ | Eurofins<br>Eurofins<br>IDT | RT-ddPCR[a] | 55 | [4] |
| N2 | F: TTACAAACATTGGCCGCAAA<br>R: GCGCGACATTCCGAAGAA<br>P: HEX/ACAATTTGCCCCCAGCGCTTCAG/BHQ1 and HEX/ACAATTTGC/ZEN/CCCCAGCGCTTCAG/IABkFQ | Eurofins<br>Eurofins<br>IDT | RT-ddPCR[a] | 55 | [4] |
| BCoV | F: CTGGAAGTTGGTGGAGTT<br>R: ATTATCGGCCTAACATACATC<br>P: /FAM/CCTTCATAT/ZEN/CTATACACATCAAGTTGTT/IABkFQ/ | IDT<br>IDT<br>IDT | RT-ddPCR[a] | 55 | [1] |
| BRSV | F: GCAATGCTGCAGGACTAGGTATAAT<br>R: ACACTGTAATTGATGACCCCATTCT<br>P: /HEX/ACCAAGACT/ZEN/TGTATGATGCTGCCAAAGCA/IABkFQ/ | IDT<br>IDT<br>IDT | RT-ddPCR[a] | 55 | [2] |
| PMMoV | F: GAGTGGTTTGACCTTAACGTTGA<br>R: TTGTCGGTTGCAATGCAAGT<br>P: FAM/CCTACCGAAGCAAATG/MGBNFQ/ | IDT<br>IDT<br>Applied Biosystems | RT-ddPCR[a] | 60 | [5] |
| HF183 | F: ATCATGAGTTCACATGTCCG<br>R: CGTTACCCCGCCTACTATCTAATG<br>P: FAM/TCCGGTAGACGATGGGGATGCGTT/MGBNFQ/ | IDT<br>IDT<br>Applied Biosystems | qPCR[b] | 60 | [6], [7] |

[a]RT-ddPCR: Primers and probes at a final concentration of 900 nM and 250 nM, respectively.
[b]qPCR: Primers and probes at a final concentration of 1 µM and 80 nM, respectively.

**Table S2** Influence of the incubation time on BCoV recovery rates and N1 and N2 concentrations

| **Sample** | **Incubation time (min) at 4°C on a rocker** | **BCoV recovery rate (%)** | **N1 (cp/L)** | **N2 (cp/L)** |
|---|---|---|---|---|
| **Green Bay DP** | 0 | 7.40 | 12137 | 6058 |
| Sept 7, 2020 | 30 | 4.79 | 14470 | 6878 |
| **Milwaukee JI** | 0 | 3.51 | 9336 | 2916 |
| Sept 9, 2020 | 30 | 2.21 | 7363 | 1949 |
| **Milwaukee SS** | 0 | 5.17 | 7423 | 2935 |
| Sept 9, 2020 | 30 | 2.66 | 6898 | 2983 |
| **Racine RAC** | 0 | 6.29 | 5666 | 5616 |
| Sept 6, 2020 | 30 | 3.37 | 5086 | 3410 |

### Supplemental Figures

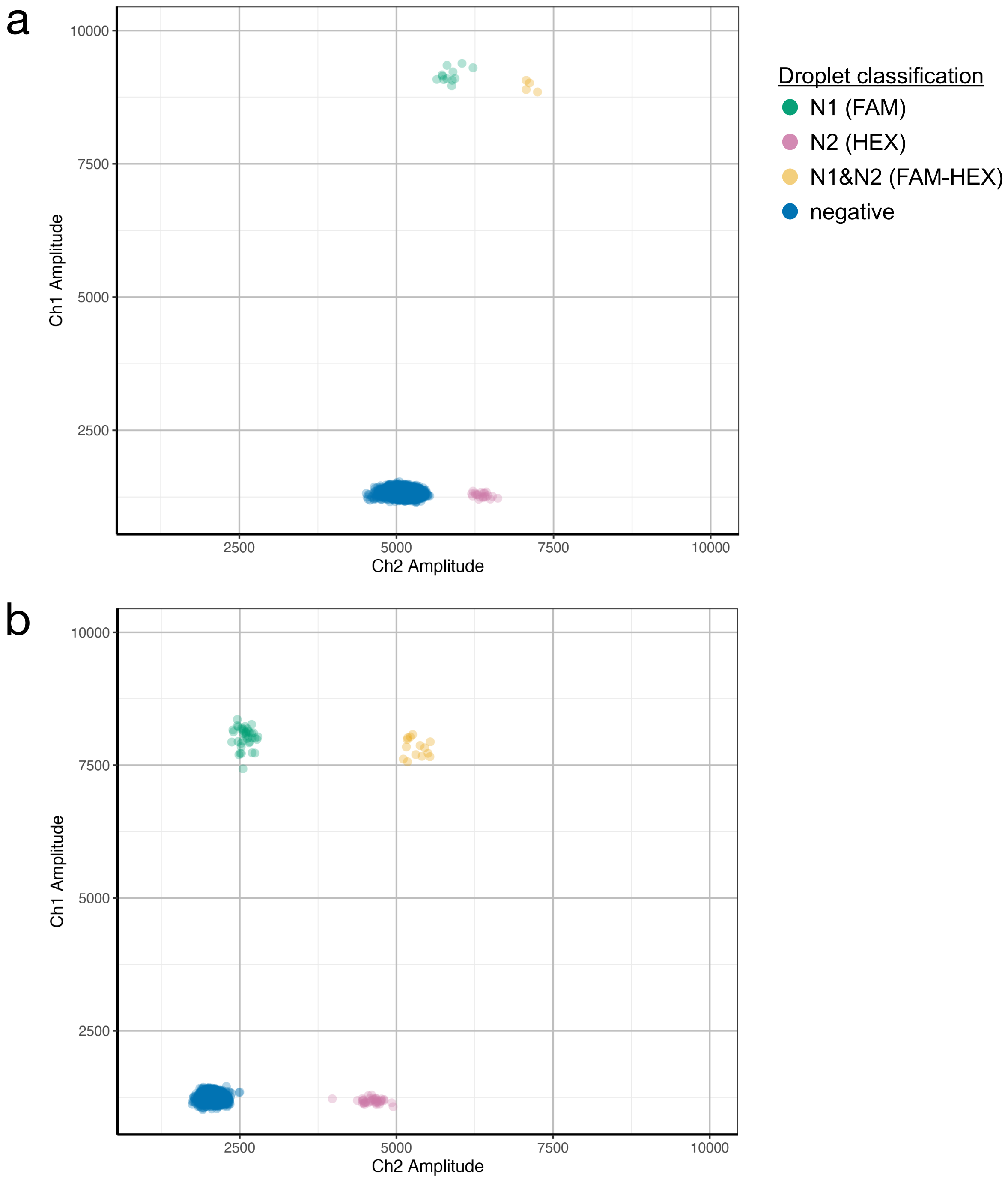


**Figure S1** Two-dimensional RT-ddPCR cluster plots of N1/N2 duplex assays using single- and double- quenched N2 probes. Channel 1 amplitude (FAM™ fluorescent reporter dye) is plotted against Channel 2 (HEX™) for (a) the N1 (5'FAM™/ZEN™/3'IB®FQ) and single-quenched N2 (5'HEX™/3'BHQ®-1) duplex assay and (b) the same N1 and double-quenched N2 (5'HEX™/ZEN™/3'IB®FQ) duplex assay. The amplitude between the negative and the N2 droplet clusters was ~1000 using the single-quenched probe and ~2500 using the double-quenched probe.

a

| | Source | Df | σ2 | σ2 rel [%] | CV [%] n=116 | F | *P* |
|---|---|---|---|---|---|---|---|
| (1) | WWTP | 5 | 0 | 0.83 | 0.48 | 12.305 | <0.001 |
| (2) | Sample | 23 | 0.03 | 52.81 | 3.86 | 25.459 | <0.001 |
| (3) | Filter | 29 | 0.01 | 22.45 | 2.52 | 5.229 | <0.001 |
| (4) | Assay (error) ε | 58 | 0.01 | 23.91 | 2.6 | | |
| | Total | 50.78 | 0.05 | 100 | | | |

$\sigma^2$ = variance component

$\sigma^2$ rel. = relative variance component (related to the overall variance)

b

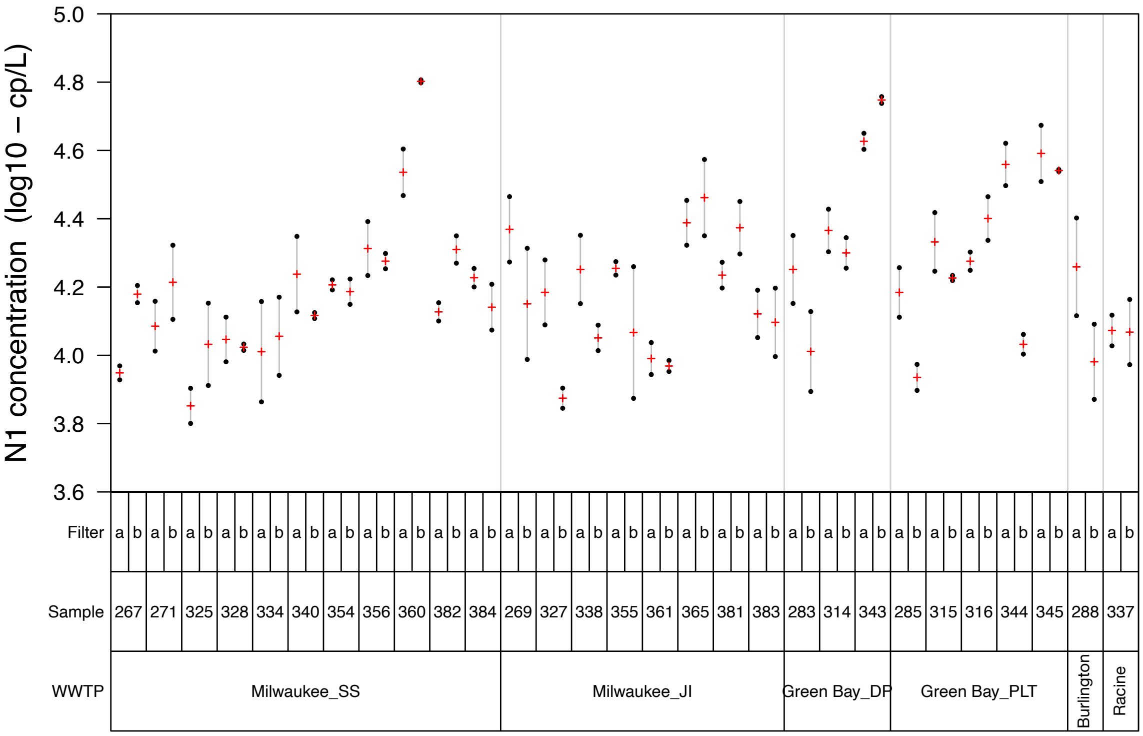


**Figure S2** (a) Nested variance component analysis results for assays, filters, samples, and WWTP. (b) Variability chart depicting the analysis. Only samples with N1 concentrations above the limit of quantification were used. Variance component analysis was performed using the function *anovaVCA* (package VCA v1.4.3) and the following command *anovaVCA* ($log_{10}$-transformed N1~WWTP/Sample/Filter), F and P values were obtained using the function *aov* and the following command *aov* ($log_{10}$-transformed N1~WWTP/Sample/Filter).

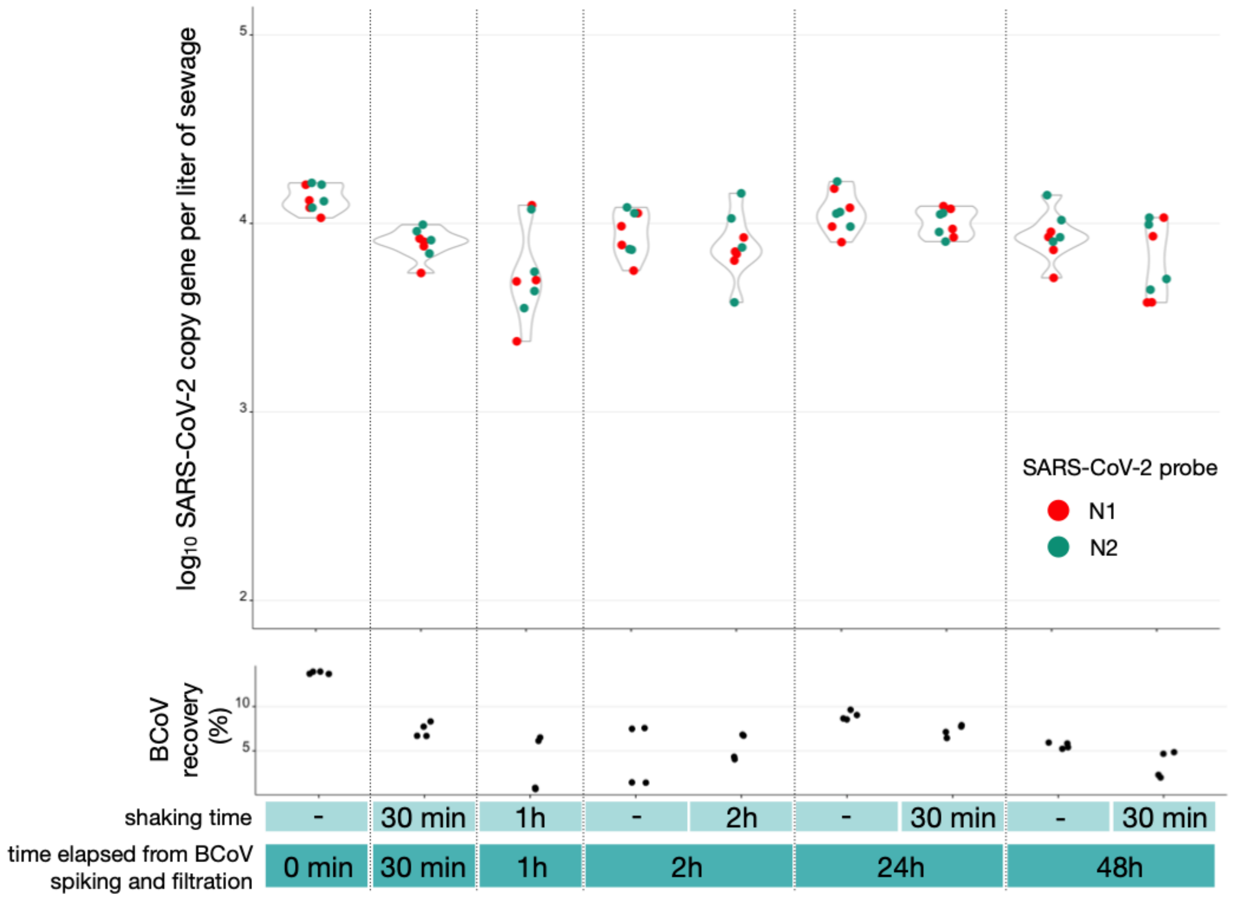


**Figure S3** Influence of BCoV incubation time (with or without shaking) on N1/N2 detection and BCoV recovery. The shaking (on rocker shaker at 4°C) was done at the beginning of the incubation.

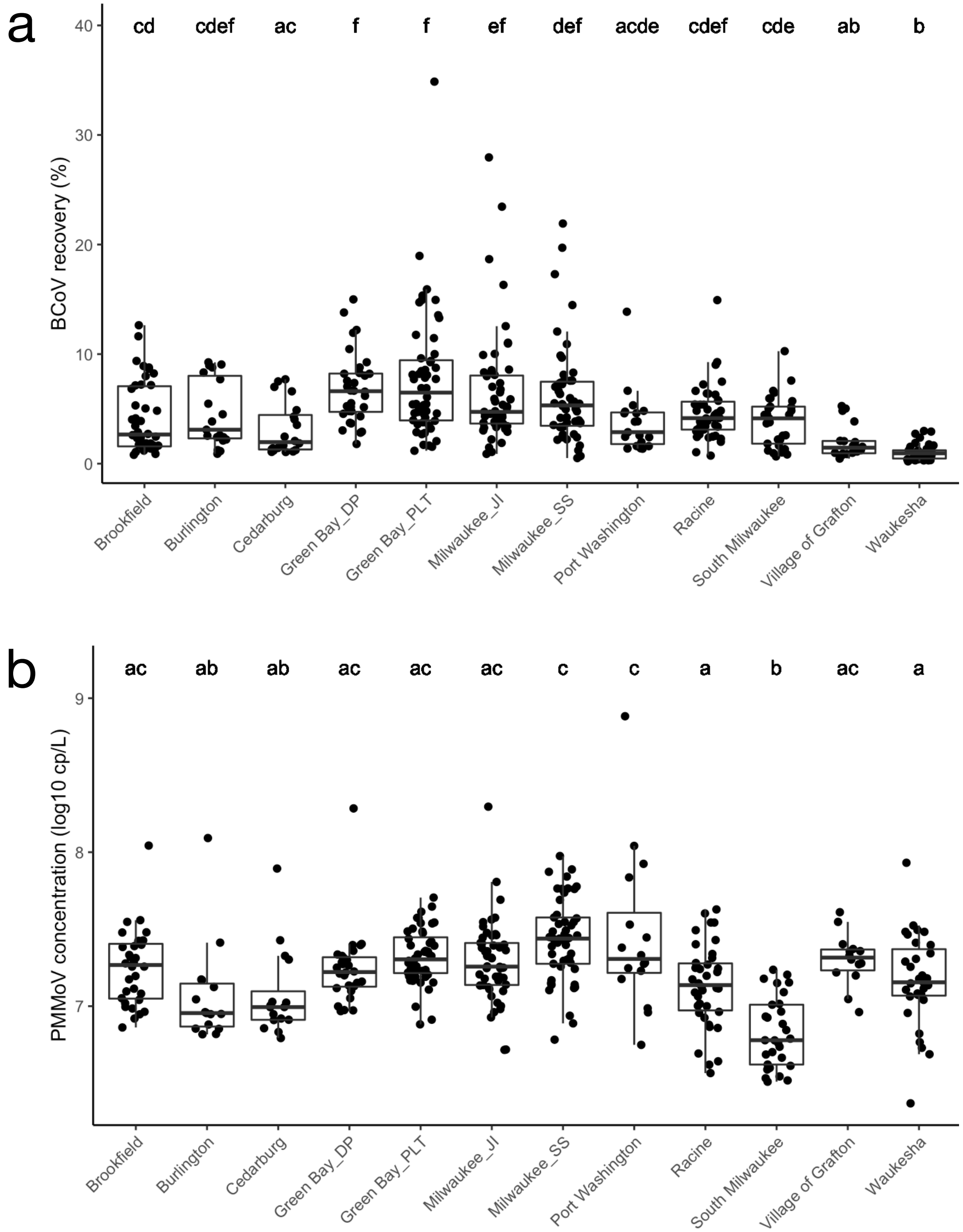


**Figure S4** Distribution of (a) BCoV recovery rates and (b) PMMoV concentrations averaged per day and per WWTP. Groups sharing the same letter are not significantly different (ANOVA, BCoV: $F(11) = 26.91$, $P < 0.001$, $log_{10}$-transformed data; PMMoV: $F(11) = 10.76$, $P < 0.001$, $log_{10}$-transformed data, Tukey-adjusted, alpha = 0.05).

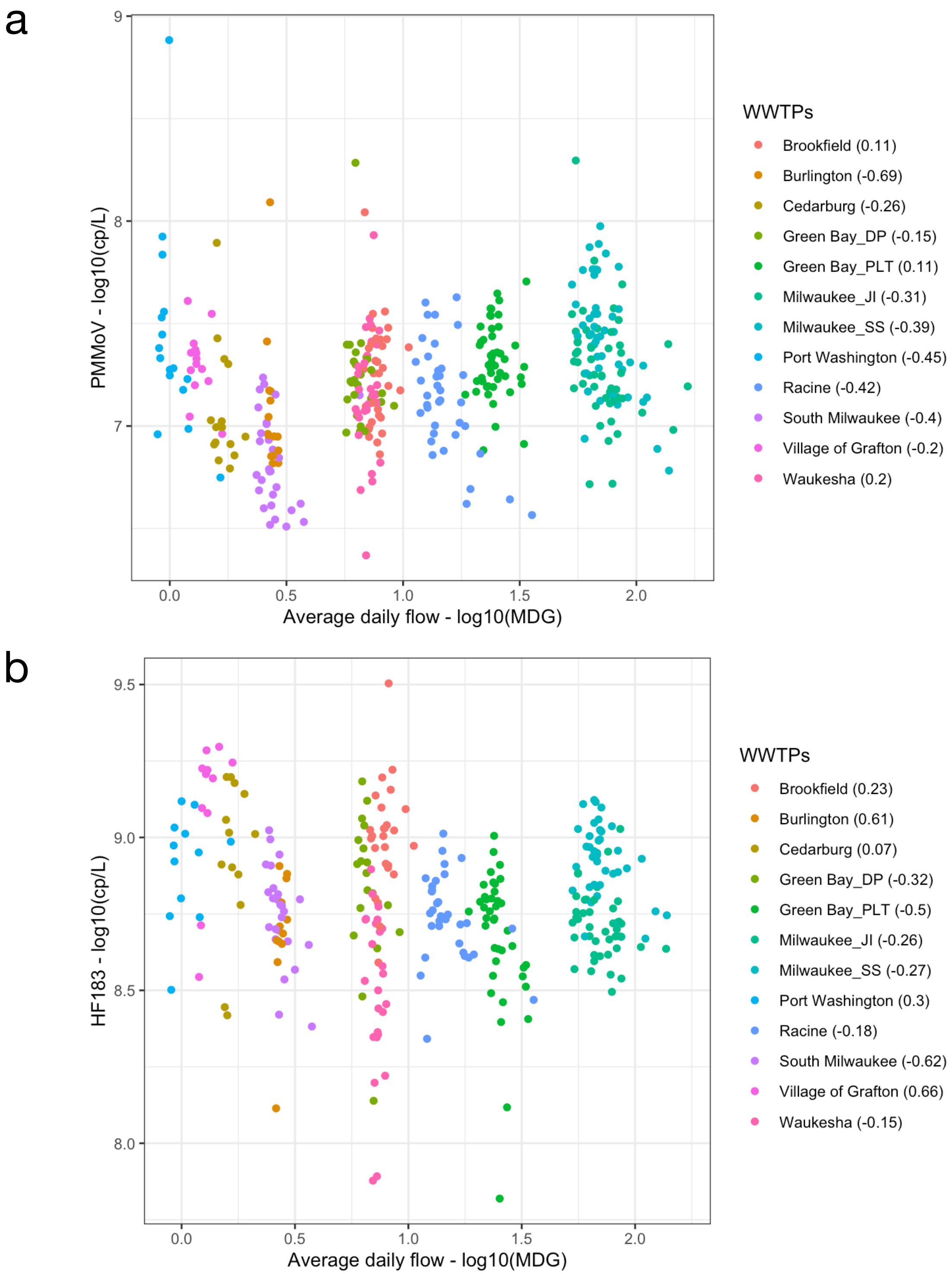


**Figure S5** Relationship between the concentrations of human markers (a) PMMoV and (b) HF183 and the average daily flow at the WWTPs. Values inside the parentheses display Spearman's rank correlation coefficients.

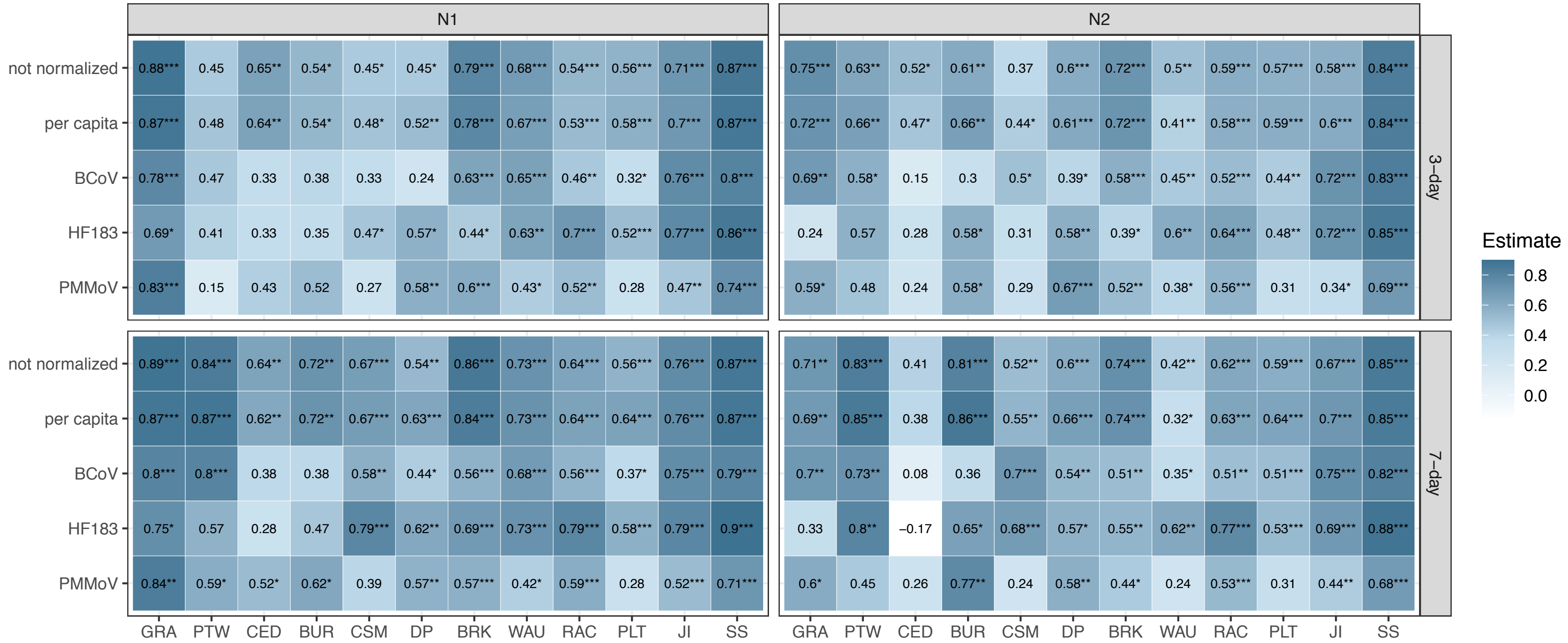


**Figure S6** Spearman's rank correlation coefficients between testing case rate per 100,000 people averaged per 3-day or 7-day and N1 and N2 concentrations in sewage without normalization or normalized to per capita in the served area, BCoV recovery rate, HF183 or PMMoV concentration. Cities are sorted from the smallest population served to the largest. Abbreviations: GRA: Grafton; PTW: Port Washington; CED: Cedarburg; BUR: Burlington; CSM: City of South Milwaukee; DP: Green Bay De Pere; BRK: Brookfield; WAU: Waukesha; RAC: Racine; PLT: Green Bay; JI: Milwaukee Jones Island; SS: Milwaukee South Shore. P-value significance level: *0.05, **0.01, ***0.001.

| Offset of positive test | | Milwaukee Jones Island | Milwaukee South Shore | Green Bay | Racine |
|---|---|---|---|---|---|
| -7 | | 0.659 | 0.838 | 0.479 | 0.687 |
| -6 | | 0.675 | 0.827 | 0.511 | 0.649 |
| -5 | Cases moved backward | 0.684 | 0.835 | 0.481 | 0.605 |
| -4 | | 0.697 | 0.855 | 0.468 | 0.636 |
| -3 | | 0.697 | 0.853 | 0.504 | 0.664 |
| -2 | | 0.730 | 0.867 | 0.512 | 0.699 |
| -1 | | 0.761 | 0.861 | 0.535 | 0.677 |
| 0 | | 0.754 | 0.869 | 0.568 | 0.688 |
| +1 | | 0.752 | 0.868 | 0.589 | 0.682 |
| +2 | | 0.758 | 0.871 | 0.559 | 0.757 |
| +3 | Cases moved forward | 0.747 | 0.869 | 0.558 | 0.735 |
| +4 | | 0.778 | 0.863 | 0.527 | 0.697 |
| +5 | | 0.797 | 0.849 | 0.596 | 0.702 |
| +6 | | 0.787 | 0.851 | 0.617 | 0.731 |
| +7 | | 0.780 | 0.839 | 0.606 | 0.683 |
| | | Positive offset 1 to 7 p<0.01 | Positive offset 1 to 7 p<0.1 | Positive offset 1 to 7 p<0.001 | Positive offset 1 to 7 p<0.01 |

**Figure S7** Spearman's rank correlations of case date (by the date of test) and SARS-CoV-2 million gene copies (MGC) per person in sewersheds. Correlations were significantly higher when cases were moved forward +1 to +7 days compared with an offset of -1 to -7 days. Since the date of test was used, this suggests that diagnosed cases are contributing to SARS-CoV-2 for several days after diagnosis, but generally not before.
